# Climate and Health policies in Argentina: obstacles and opportunities from stakeholders’ perspectives

**DOI:** 10.1101/2025.08.27.25334531

**Authors:** Luciana Beatriz Castronuovo, Cinthia Shammah, Clara Agustina Trebucq, Andrea Hurtado, Damián Verzeñassi, Milena Sergeeva

## Abstract

In Argentina, climate change poses significant challenges for public health, affecting heatwave incidence, vector-borne disease distribution, and healthcare infrastructure resilience. Despite being the first Latin American country to explicitly integrate health into its Nationally Determined Contributions (NDCs) and establishing a National Strategy for Climate and Health, the extent to which these frameworks translate into tangible action remains unclear.

A total of 31 key informants from national and subnational governments, research institutes, healthcare facilities, and international organizations were interviewed using a semi-structured format. Data were analyzed using an operational framework for building climate-resilient and low-carbon health systems, which was refined inductively to identify core themes and categories based on interviews and a workshop with key stakeholders.

Findings reveal notable progress in governance, including the creation of intersectoral spaces and policy instruments. However, significant barriers persist: limited political will at higher levels of government, insufficient local implementation capacity, fragmented health information systems, and a lack of sustained funding—particularly for adaptation. At the same time, Argentina’s federal structure allows provinces and municipalities to advance climate and health initiatives independently, which may be perceived as a barrier or as an opportunity depending on the context. Technical teams and personal networks often maintain progress during periods of fluctuating political support.

Argentina’s experience underscores both the potential and the complexity of integrating climate and health agendas. By identifying key facilitators and obstacles, this study offers valuable lessons for other Latin American countries. Strengthening data interoperability, expanding local implementation capacity, securing consistent funding, raising awareness among the general population, health care professionals and other relevant stakeholders to build a common consensus on the urgency of tackling climate change will be critical for building a truly climate-resilient health system.

## Introduction

Climate change affects human health through both direct pathways (e.g., heat-related illness, respiratory and cardiovascular mortality, hospitalizations) and indirect pathways (e.g., disruptions to health services, increased transmission of waterborne diseases, infrastructure damage) (1). The magnitude of these impacts is mediated by social and economic factors, often disproportionately affecting vulnerable populations. From a global perspective, low-and middle-income countries (LMICs) are particularly susceptible to climate-related hazards (2,3). While some studies have examined the resilience of health systems in Africa (4–6), Asia (7), and Latin America (4), evidence remains limited—especially in Central and South America (8). This region is among the most urbanized in the world and is characterized by high levels of inequality. According to the latest Intergovernmental Panel on Climate Change (IPCC) report, it is already experiencing a growing frequency and severity of extreme events, including heatwaves and droughts (9).

Argentina presents a particularly relevant case: it was the first country to include health in its Nationally Determined Contributions (NDCs) in 2020 (10,11). The country has also made institutional advances, such as the enactment of Law 27,520 on Minimum Standards for Adaptation and Mitigation to Climate Change in 2019 (12), the release of two National Plans for Climate Change Adaptation and Mitigation (in 2019 and 2022) (13,14), and the development of a National Strategy on Health and Climate Change in 2023 (15). These policies were developed under two different administrations that both maintained a political commitment to the climate-health nexus. This study was conducted during a period of political transition, following the inauguration of President Javier Milei in December 2023.

Argentina is expected to face increasing health threats due to the climate crisis (13). For instance, the number of heatwave days is projected to rise (16,17), and the distribution and seasonality of vector-borne diseases may shift due to changes in temperature and precipitation patterns (18).

Building climate-resilient health systems is a global priority. The World Health Organization’s (WHO) 2023 Framework for Building Climate-Resilient and Low-Carbon Health Systems offers a comprehensive theoretical model for assessing how health systems respond to climate challenges (19). This framework encompasses both adaptation and mitigation measures and has been applied in diverse settings, with adaptations to accommodate specific health system structures and regional needs (20–22).

In light of the need to strengthen Argentina’s health system resilience, this study—the first comprehensive analysis of climate and health policy in the country—applies the WHO framework to examine existing initiatives and identify key barriers and opportunities for effective implementation.

## Methods

### Sample

We conducted 31 in-depth interviews with key informants involved in climate and health policy in Argentina (Table 1). The recruitment period started on 1 May 2024 and ended on 9 August 2024. The interviews were carried out between May and August 2024, during a period of political transition. A total of 26 semi-structured interviews were conducted—mostly one-on-one, although a few involved more than one participant. We used purposive sampling to identify stakeholders involved in developing the National Strategy on Health and Climate Change, and complemented this with snowball sampling. Hospital-level stakeholders were identified in collaboration with Health Care Without Harm (23). Most interviews were conducted with national-level actors, though we also included provincial-level informants. The province of Neuquén was selected due to its participation in a Green Climate Fund-supported Readiness Program (24) and was highly recommended by multiple informants.

**Table 1:**
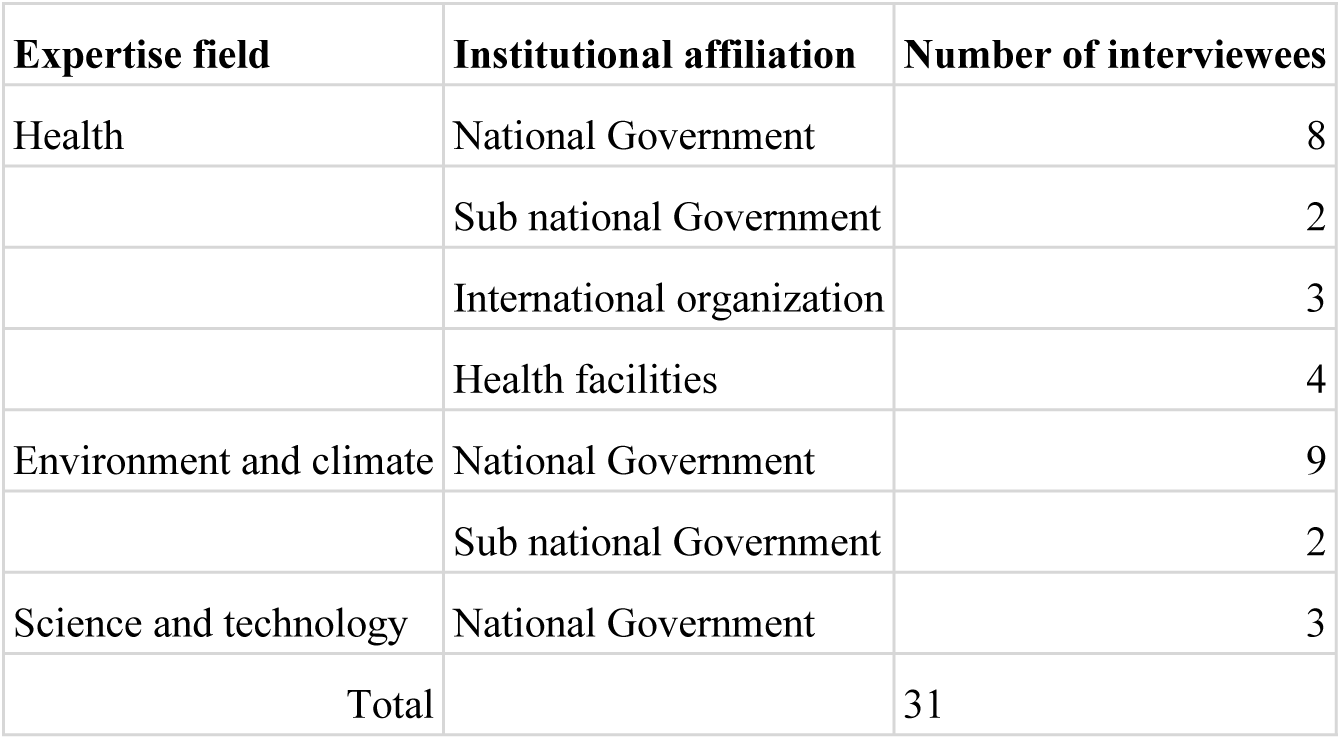
Sample description.

| Expertise field | Institutional affiliation | Number of interviewees |
| --- | --- | --- |
| Health | National Government | 8 |
|  | Sub national Government | 2 |
|  | International organization | 3 |
|  | Health facilities | 4 |
| Environment and climate | National Government | 9 |
|  | Sub national Government | 2 |
| Science and technology | National Government | 3 |
| Total |  | 31 |

### Data Collection

Interviews were conducted by experienced qualitative researchers following best practices to ensure data reliability (25). Each session involved at least two researchers, and a preparatory document summarizing the interviewee’s role was developed in advance. The interview guide was adapted based on the type of stakeholder—national or local actors, representatives of international organizations, or hospital staff—and tailored to each participant’s background. The guide was pre-tested with key informants. Field notes were shared with the research team to enhance reflexivity (26). We achieved a high participation rate, with only three individuals declining.

Relevant official documents and policies mentioned by interviewees were also reviewed to contextualize findings (27,28).

Following the interviews, a workshop was held with more than 20 key informants, including representatives from government, academia, and civil society. The analytical dimensions and their definitions were discussed and agreed upon by participants. Facilitated group discussions were conducted, and detailed minutes were developed

### Data Analysis

We used the WHO 2023 operational framework as our theoretical foundation to assess the resilience of Argentina’s health system (19). An inductive approach was then applied to refine and adapt the framework’s dimensions based on the empirical data. One researcher performed the initial coding, generating categories and themes, which were then reviewed and validated by two additional researchers to ensure consistency and rigor (29).

We made some adaptations to the WHO framework. The dimensions "Service Delivery" and "Essential Medical Products and Technologies" were merged into a broader category, "Adaptation and Mitigation in Health Facilities," due to limited data saturation on specific medical technologies. Similarly, we broadened the "Health Workforce" dimension to include a wider array of actors, including ministries and NGOs. Additionally, we chose to emphasize capacity building and awareness raising as key components of our approach. While the WHO framework provided valuable structure, we applied flexibility to allow the data to shape the analysis rather than imposing predefined categories.

### Ethics

All participants provided written informed consent via DocuSign prior to participation, and signed forms were securely stored by the research team. The study received ethical approval from the Iniciativa y Reflexión Bioética (IRB) at both the national level and in the Province of Buenos Aires. The IRB in the Province of Neuquén determined that the study did not fall within its scope, as no personal health data were collected.

## Results

For each dimension —1. Leadership and governance; 2. Capacity building and awareness of the health sector; 3. Health Information Systems; 4. Financing; 5. Adaptation and mitigation in health facilities— we identified different themes (Table 2) as well as key barriers and opportunities (Table 3).

**Table 2.** Dimensions and themes.

| Dimension | Theme |
| --- | --- |
| Governance and leadership | Intersectoral articulation |
|  | Technical Networks |
|  | Lack of communication |
|  | Political will as an enabler |
|  | Gap between governance and concrete actions in territories |
|  | Political will as a challenge |
|  | Federalism (opportunity/challenge) |
| Capacity building and awareness of the health sector | High expertise and knowledge |
|  | Shared commitment |
|  | High turnover of staff as an obstacle |
|  | Difficulty in perceiving the climate-health connection |
| Health Information Systems | Difficulty in sharing data and interoperability |
|  | Challenges in the generation of climate and health data |
| Funding | Funding tends to prioritize climate change mitigation over adaptation |
|  | Readiness Project |
|  | International funding dependence |
| Adaptation and mitigation in health facilities | Lack of comprehensive approach |
|  | Emergency approach |

**Table 3.**
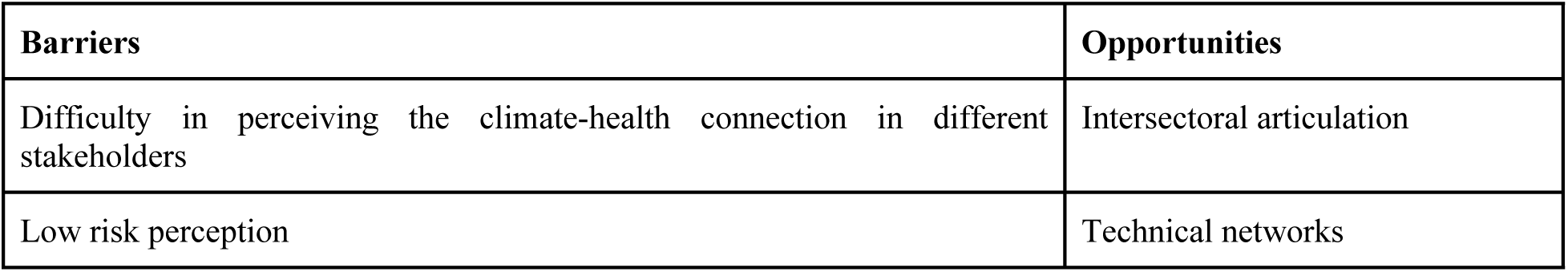
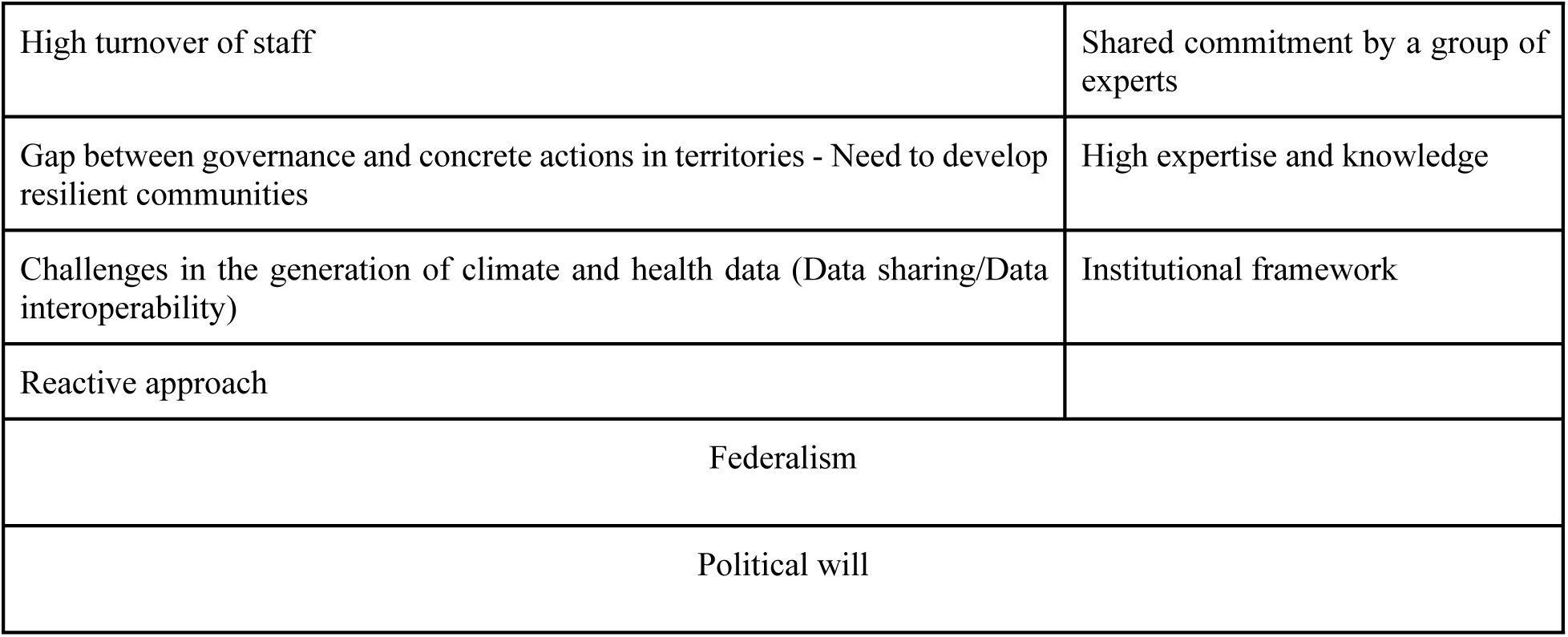
Barriers and opportunities identified by interviewees.

### 1) Leadership and governance

The interviewees identified an initial set of plans (13–15,30–32) developed under the National Cabinet for Climate Change (NGCC), established in 2019, that began to address the relationship between climate and health (Figure 1). One of the main instruments highlighted was the 2019 National Action Plan for Health and Climate Change (30), which outlined specific adaptation goals for the health sector. These included strengthening the health system’s capacity to respond to cold and heat waves, floods, and mosquito-borne diseases, as well as improving the resilience of healthcare facilities to extreme weather events (Figure 1).

**Fig. 1:**
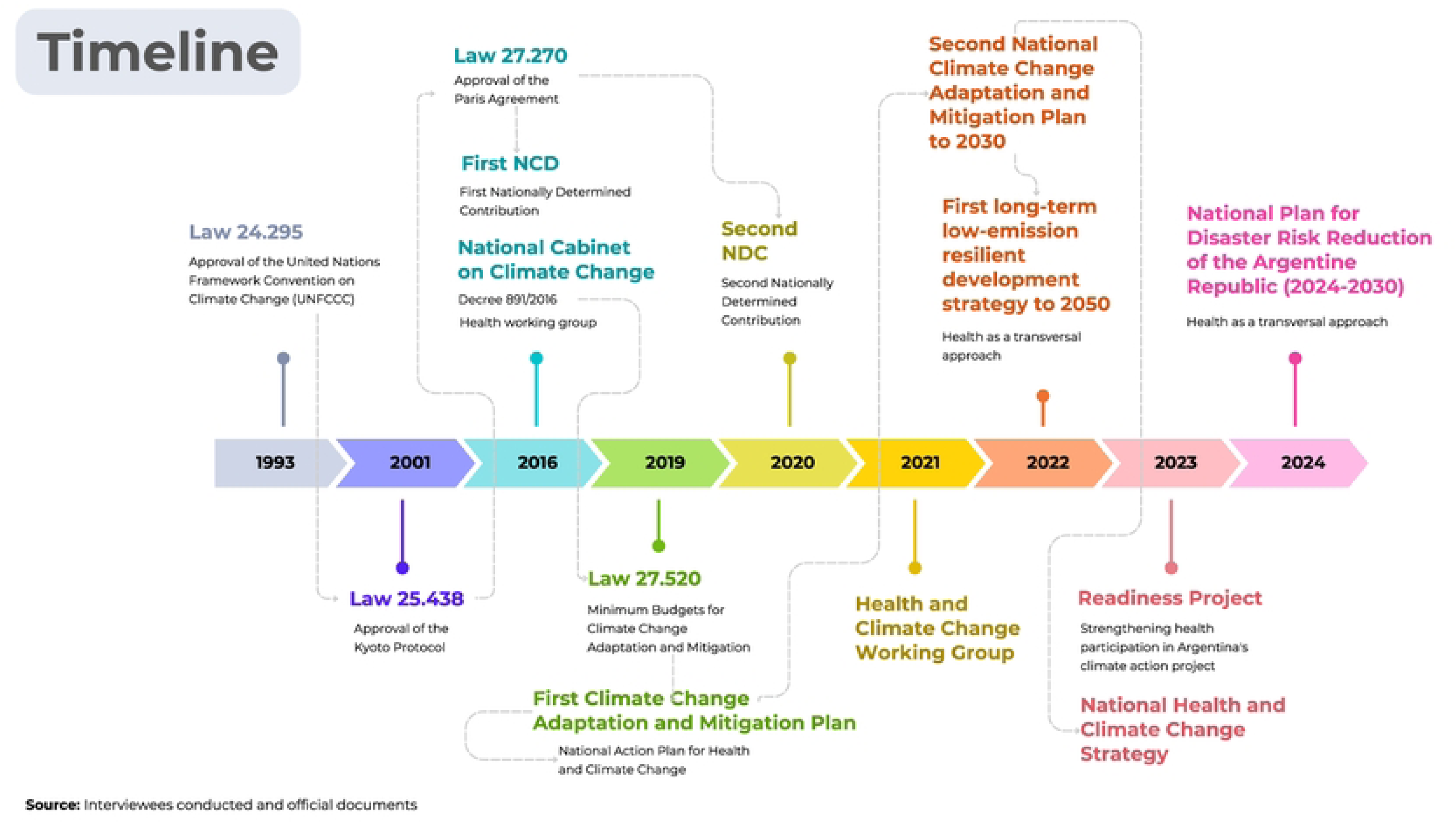
Governance framework timeline

Different interviewees mentioned difficulties in communication between different sectors and levels of government. To close this gap different governance spaces were created where different stakeholders could meet and work together intersectionality. Interviewees mentioned that the COVID experience highlighted the urgent need to develop such spaces. One of the persons involved in creating one of these spaces, which is still functional to this day, highlighted:

> *“the first, I don’t know, three, two meetings, it was like they were Greeks and Trojans. Nobody had ever met, they did not know what”* (Interviewee 3. Health sector, National Government)

At the national and subnational governance level, progress is evident in the development of regulations, plans and strategies. Some successful examples of this intersectoral collaboration include: risk maps and early warning systems which were the product of people working together from different sectors. As an interviewee from the health sector mentioned:

> *“We have an early warning system for extreme heat and cold temperatures operated by the meteorological service, which informs the health system and the health system generates risk communication (…) This was funded by the Ministry of Health. (Interviewee 2. Health sector, National Government)*

Despite these advances, many interviewees agreed that translating regulatory and institutional frameworks into concrete actions is a challenging task. As one informant noted, “*the great challenge has always been how to translate what we are writing on paper, which can be beautiful and important for all of us, into reality, that is: how we can make it happen” (Interviewee 8. Environmental and Climate sector, National Government)*

The interviewees also point out that in a context where political will at the national level is withdrawn, federalism becomes an opportunity, as each province, municipality and territory have a certain autonomy. As one interviewee highlighted, “*regardless of national guidelines, the municipalities have not stopped their actions, in fact many more have advanced their efforts” (Interviewee 30. Environmental and climate sector, subnational level)*

### 2) Capacity building and awareness of the health sector

All interviewees agreed that Argentina has a wealth of professionals with extensive expertise and knowledge in climate and health issues. Many of those interviewed, along with their colleagues, adopt a proactive approach in their daily work, taking on tasks beyond their formal responsibilities to ensure that climate and health issues are increasingly integrated into their respective agendas. For example, when the National Climate Change Cabinet was created in 2016 and established which government sectors should participate, the Ministry of Health was not included. However, professionals committed to the issue sought a way to join the cabinet. As one health sector representative involved in this process explained, they "*got involved thanks to those articles that say “if necessary, others may be summoned” (…) later on, to the surprise of those who were leading the Cabinet at that time, we were one of the few ministries that developed its Sectoral Plan*" (Interviewee 2. Health sector, National Government)

Professionals committed to climate and health also form strong connections with other dedicated professionals, particularly within technical teams, and advocate for policy change in an effective way. Progress on climate and health has largely been driven by highly committed individuals or small groups. A former representative of the Environmental sector pointed out, "*Much of what is happening is thanks to the efforts of the technical teams*” (Interviewee 13. Environmental and Climate sector, National Government)

An interviewee from a national research institute observed that when political will shifts and no longer supports climate and health initiatives, actions can still be sustained through technical and personal networks. As another interviewee explained, these relationships enable continued collaboration and exchange when higher levels of political leadership withdraw support: *“the interest and appropriation of the agenda by these local agents, which seems to me to be the possibility of sustaining these discussions (…) this is the great hope in the absence of concrete action on the part of the national government”. (Interviewee 8. Environmental and Climate sector, National Government)*

Regarding the awareness of healthcare personnel at the hospital level, interviewees consistently highlighted that physicians are not sufficiently sensitized to the climate challenge. As one interviewee mentioned: *"it is as if they do not have it incorporated, as if they are not aware of it. (…) They don’t take it as something that impacts health." (Interviewee 26, Health sector, Health facility).* There is a lack of clear recognition of the impact of climate change on health, even when these effects manifest in clinical consultations. For instance, while physicians routinely treat cases of dengue fever, they do not necessarily establish a connection between the increasing incidence of the disease and climate change. This disengagement reflects a prevailing perception that environmental health is not within their scope of responsibility. Furthermore, hospitals generally adopt a treatment-focused approach, often at the expense of preventive measures. This tendency aligns with the institutional mandate of general hospitals, which prioritize treatment, while primary healthcare facilities are more actively involved in prevention and health promotion efforts. In short, the healthcare workforce is not sensitized to climate and only some of them receive training in environmental health.

### 3) Health Information Systems

There has been relevant collaboration to produce data between institutions, such as the National Meteorological Service, the National Commission for Space Activities and some universities. In the area of information systems, key developments include the creation of early warning systems, risk maps for vector-borne diseases such as dengue, and the stated intention—reflected in official documents and regulations—to establish a climate change and health observatory. At the same time, health information systems face a series of difficulties.

A critical challenge is the generation of climate and health data. Interviewees agree that data reflecting the impact of climate change on health is difficult to conceptualize as the signals are often subtle or indirect. This complicates the use of morbidity data to accurately reflect climate-related health impacts, compromising the effectiveness of epidemiological surveillance. As one interviewee from the national health sector explained,*“(…) Heat waves or cold snaps have a looser case definition [than other diseases] that can encompass a fairly broad and very non-specific range of signs and signals”(Interviewee 18. Health sector, National government)*

Additionally, mortality data—crucial for assessing the effects of extreme weather events—often takes time to consolidate, delaying timely decision-making. Likewise, interviewees mentioned difficulties in sharing data without a defined protocol for conducting this process. Furthermore, information on the health impacts of climate change that is indeed developed is scattered across various sources and formats, hindering data interoperability. The lack of systematization of this data, combined with a federal system and state fragmentation, adds to the challenge of integrating and utilizing this information effectively. As one interviewee from the environmental and climate sector explained, *“honestly, there is a difficulty in accessing information, especially because, well, the national health system is quite compartmentalized, isn’t it? You have national, provincial, municipal, public, private. It is quite diverse”(Interviewee 13. Environmental and Climate sector, National Government)*.

Some interviewees commented that the data generated is not always shared. This may be due to the complication of data ownership, unwillingness or reluctance to share some data considered sensitive. This situation was clearly illustrated by a national-level representative from the environmental and climate sector, who noted: *“we are still, apart from being egocentric, we are jealous of our data, if it is generated by my ministry, it belongs to my ministry, I want to publish it, make the book, the minutes, everything” (Interviewee 1. Environmental and Climate sector, National Government)*

While the effects of climate change are visible in local communities, the data often remains too abstract, general, and disconnected from everyday experiences, making it hard for people to grasp. Several interviewees stressed the importance of *bringing the data down to earth*—making it more tangible, understandable and usable for broader audiences. One of the interviewees mentioned:*“I say a lot to my colleagues who work on climate change, even we don’t know what "[to limit global warming to]1.5ª" is. So you can’t pretend that this generates a demand for public policy among the people”(Interviewee 16, Environmental and climate sector, National Government)*

At the same time, data provided by local territories is not sufficiently integrated into political decision-making processes, reflecting a lack of effective bottom-up coordination. Data is often based primarily on top-down scientific-technical systems.

Some good examples of data sharing and integration identified are the warning system for heat and cold waves developed by the National Meteorological Service in coordination with academia and the National Ministry of Health. In addition, there is effective communication between the Epidemiology Department of the National Ministry of Health and the National Meteorological Service. The latter informs the Ministry about upcoming heat waves, allowing them to organize a preventive response.

### 4) Financing

The primary sources of funding for policies can be categorized into two main streams: international funding and national budget allocations. Interviewees recognized that those projects that address climate change will have a better chance to attract International funding. This is a motivation for some areas to include climate change in their projects. An example of international financing is the Green Climate Fund-supported Readiness project, to strengthen climate and health plans of three provinces: Neuquén, Tucuman and Misiones. The aim was for these provinces to be beacons for each region, with their best practices serving as models that could be scaled to other provinces. An interviewee from an international organization explained:

> *“Neuquén (an Argentinean province) has become the motivator and the one that explains or tells them about the good practices they have had, for example within the framework of readiness, and they are motivating the provinces of the Patagonian region” (Interviewee 23, International Organization)*

Several interviewees agreed that Argentina’s governance structure—particularly the development of its Climate and Health Strategy and the inclusion of health in its Nationally Determined Contributions (NDCs)—played a key role in enhancing the country’s eligibility for the Green Climate Fund Readiness Project.

The financing of climate and health policies in Argentina is characterized by a reliance on external funding for policy development and implementation. The lack of dedicated state resources in this area restricts the capacity of teams to act effectively, creating a dependency on international cooperation. As one interviewee from the environmental and Climate sector pointed out:*“In Argentina in particular, but also to other countries in general, always developing countries in particular, we are looking for financing for, because we need it for absolutely everything or for many things and everything related to the environment and climate change, the truth is that it depends almost 100% on international financing” (Interviewee 13. Environmental and Climate sector, National Government)*.

Several interviewees noted that international funding tends to prioritize climate change mitigation over adaptation, even though developing countries, like Argentina, have a greater need for support in adaptation efforts. One interviewee highlights that *“unfortunately, at a global level, those who give money, (…) those who are responsible for financing these issues, are not disbursing their fair share for adaptation, the adaptation funds are pretty…empty.” (Interviewee 13. Environmental and Climate sector, National Government)*

Although some interviewees have mentioned the complementarity of the adaptation and mitigation axes, they consider them to be separate in practice and consider that mitigation should not be the focus.

### 5) Adaptation and mitigation in health facilities

Interviewees agree that the Argentine health sector currently approaches the impacts of climate change from an emergency approach, reducing the comprehensive perspective of integrated risk management. Interviewees argue that climate change policies are delayed as priority is given to other things which are perceived to be more urgent. As one expert highlighted, *“It can’t be that at the last minute you call health to go out to support. It’s the same with emergencies. It is always like that. So, “no, well, we don’t know what to do” and then the doctors and nurses and the whole health system, professionals and non-professionals, are there to see what they can do with the remains of what is left here, all chopped up, fallen*” (Interviewee 3. Health sector, National Government)

The response to the impact of climate change on health becomes reactive: it is deployed once a disaster and emergency occurs (for example, when there is a disproportionate increase in dengue cases), but the necessary preventive actions to avoid the impact of climate change on communities are not considered. Interviewees emphasize the need to incorporate a comprehensive approach and work from a preventive perspective.

### 11. Health sector, National Government)

Once the problems are hitting the territories, a possible response is deployed. The interviewees mention that this situation must be understood in a context where different state areas suffer from different economic and management constraints and high staff turnover. As one expert noted, “*[They] increase personnel only when you are in an outbreak and therefore it is a personnel that is trained at that time, the outbreak ends and the personnel is precarized and their contracts are cut and therefore the following year you find yourself with new personnel or personnel that do not exist” (Interviewee 5. Environmental and Climate sector, National Government)*

Similarly, among health professionals, the treatment of patients prevails over concern for the impact of climate change on health. In cases where actions connected with climate where implemented in health care facilities were not the consequence of an institutional framework but of highly committed individuals inside the organization. For example, one private hospital has developed a comprehensive crisis, catastrophe, and climate change adaptation plan tailored to its specific vulnerabilities, such as floods, droughts, and high winds. This plan has evolved over the years, integrating environmental variables to enhance the hospital’s response capacity to extreme weather events. Regular evacuation drills are conducted, and staff training focuses on emergency preparedness. This initiative, driven by a committed team and backed by institutional support, exists independently of any state-level policy or plan. According to the interviewee from this hospital: *"we started to see that we had to work with the community because if something really happens, people run to the hospitals. So, what was born as a ‘multiple victim care plan’ started to expand a little bit more, today it is a crisis, catastrophe and climate change adaptation plan" (Interviewee 29. Health sector, Health facility)*

## Discussion

This is the first study to analyze climate and health-related policies in Argentina. Our findings highlight Argentina’s pioneering role as the first country to include health in its Nationally Determined Contributions (NDCs) and its significant progress in climate and health governance, as evidenced by various plans and strategies. The importance of these governance structures becomes especially clear during government transitions.

Progress in governance and successful hospital-level initiatives have largely been driven by political will and a group of technically trained, committed stakeholders. As such, engaging key actors remains essential. Until now, the implementation of climate and health policies in Argentina has followed a predominantly top-down approach. Moving forward, it is crucial to incorporate more bottom-up strategies, prioritizing community voices and implementing concrete actions at the local level. Continued efforts to generate local-level data on the health impacts of climate change are vital for guiding these actions.

Argentina’s governance progress can be characterized as a process of "layering," where successive initiatives build upon each other, generating cumulative and mutually reinforcing effects. Strengthening governance structures is essential to effectively address the challenges of the global climate crisis (2,33). The creation of multi-stakeholder spaces reflects an increased awareness of the need for cross-sectoral policymaking. While some interviewees reported difficulties in coordination across government sectors—a finding echoed in the literature (4,34,35)—the COVID-19 pandemic served as a catalyst for intersectoral collaboration (3,36), resulting in the establishment of new coordination mechanisms. If effectively implemented, these governance structures can help overcome longstanding barriers.

However, a notable gap persists between formal governance structures—such as regulations, plans, and strategies—and the actual implementation of policies. This gap is evident in other high-, low-, and middle-income countries as well (4,37–39).

Although evidence on the climate resilience of health systems is limited—especially in South America—findings from previous studies align with ours. Both prior research and our own results emphasizes the need to raise awareness about the impact of climate change on health among the general population, healthcare professionals, and sectors of government that have yet to prioritize the climate-health nexus (4,38,40,41).

While best practices have emerged in some health facilities and a core group of highly engaged health workers has been identified, widespread awareness among healthcare professionals remains low (42). This highlights the urgent need for targeted training on climate and health for the healthcare workforce, especially given the strain on health systems in low- and middle-income countries (LMICs) (3) A paradigm shift is needed to focus on building a preventive and comprehensive response to climate change. Often, the health impacts of climate change are deprioritized until a crisis emerges. Health professionals tend to focus on more immediate concerns, pushing climate-related issues down the agenda (42).

Moreover, it is important to highlight that initiatives like the Readiness project can help scale successful practices nationwide. For example, in June 2025, a meeting between Tucumán—a province that previously participated in the Readiness project—and four other northwestern provinces aimed to share results and initiate joint planning to expand successful outcomes (44).

Many individuals involved in developing Argentinean policies form a network of technically skilled and committed actors. These networks, recognized in other studies as key facilitators (45), have contributed to the development of early warning systems, supported by institutional frameworks. Cooperation between meteorological services and health actors is seen as a critical facilitator (37).

The availability of local data is vital for raising awareness among stakeholders about the health impacts of climate change. Yet studies consistently report difficulties in data production, sharing, and integration (22,39,40). Despite these obstacles, Argentina can leverage regional experiences (46) to further develop its Climate and Health Observatory, a component of the National Climate and Health Strategy.

Challenges in data integration reflect a siloed approach that hampers effective policy implementation and highlights persistent incoherencies, even in the presence of governance structures.

During the study period, a new government took office in Argentina, prompting concerns about the effects of political transitions on policy continuity. Political will and polarization around climate change emerged as key factors shaping the policy landscape. Although existing policies can support continuity, shifting government priorities may compromise climate and health initiatives, including funding and institutional support (4,47). In this context, community involvement becomes even more crucial for sustaining public and preventive health services (2).

Interviewees noted that Argentina’s federal structure can be both a barrier and an opportunity. While state fragmentation can hinder progress, in the absence of national support, federalism can empower subnational actors to implement localized climate and health policies. Provinces and municipalities maintain autonomy to advance such policies independently. The Readiness project offers a model for subnational policy design. Despite national-level obstacles, the efforts of technical teams, their networks, and local data development remain essential to strengthening local health systems (48).

To overcome the challenges identified in this study, several actions are recommended. First, documenting and sharing best practices—such as early warning systems and green hospitals—can encourage wider adoption. Second, the generation of local data must be reinforced, and a Climate and Health Observatory should be established to consolidate initiatives and monitor progress. At the advocacy level, governance structures should be monitored and strengthened for sustained implementation. Subnational collaboration should be prioritized, and communication strategies should aim to raise awareness among decision-makers. Building alliances with environmental stakeholders and developing a unified narrative can amplify the message on climate and health. In short, developing a climate-resilient health system must be seen not as optional, but as an urgent necessity.

### Strengths and Limitations

This is the first study in Argentina to analyze climate and health policies. The insights from interviewed actors offer valuable perspectives on the complexities of a changing political environment. After successive administrations supporting climate policy, political will has now diminished. By identifying the facilitators and barriers to climate and health policies in this shifting context (49), the study provides important input for policymakers across the region. However, there are limitations. Most interviews were conducted with national-level informants and civil servants. Future research should expand to include more provincial and municipal perspectives. Additionally, impact evaluations and vulnerability assessments across different regions and sectors (e.g., transport, energy) are needed.

## Conclusions

Argentina has made strides toward building a climate-resilient health system, particularly by developing governance structures such as national strategies, plans, and its inclusion of health in its NDC. These steps have enabled international funding and fostered cross-sectoral collaboration, breaking down silos—as seen in the development of early warning systems. The role of committed individuals across sectors (health facilities, government, academia) has also been critical.

However, significant challenges remain. First, an interoperable, accessible health information system using local evidence is needed to sensitize stakeholders and inform context-specific policies. Second, there is a need to move beyond reactive approaches and recognize the urgency of climate and health policy—a challenge in a resource-constrained setting. Finally, it is vital to raise awareness among communities, health professionals and decision-makers about the relevance of climate change to health.

## Data Availability

The data generated and analyzed during this study are not publicly available due to ethical restrictions. The interviews conducted include not only participants' names but also details about their professional backgrounds, institutional affiliations, and policy roles, which make them potentially identifiable. Given the sensitive nature of the information shared and the commitments made to participants regarding confidentiality, we are unable to share the full transcripts or audio recordings. Requests for access to anonymized excerpts of the data may be considered on a case-by-case basis and should be directed to the Ethics Committee Iniciativa y Reflexión Bioética (IRB).

## Acknowledgments

We would like to thank all the participants of the study and the research, advocacy and communication areas of FIC Argentina who provided insights to this research. We also would like to thank Mariana Saidón and other members of the Área de Ambiente y Política-Escuela de Política y Gobierno-Universidad Nacional de San Martín. We also would like to thank Robert Marten and Idil Shek Mohamed for the relevant contributions to the manuscript.

